# Mechanistic 5’UTR Variant Scoring Expands Rare Variant Discovery in the UK Biobank

**DOI:** 10.64898/2026.09.09.26362607

**Authors:** Matthieu Chaldebas, Khoren Ponsin, Haralambos A. Mourelatos, Yoann Seeleuthner, Clément Conil, Jonathan Bohlen, Jean-Laurent Casanova, Peng Zhang, Aurélie Cobat

## Abstract

The 5′ untranslated region (5′UTR) regulates protein output through upstream open reading frames (uORFs) and Kozak context, yet most deleteriousness scores rely heavily on evolutionary conservation of its nucleotide positions. Using 5ULTRA, a machine-learning classifier trained on 5′UTR regulatory biology, we annotated rare and low-frequency 5′UTR variants in 408,423 UK Biobank participants. We tested gene-level associations for all 59 quantitative blood-count and serum biochemistry traits. We identified 58 genome-wide significant gene-phenotype associations and CADD 38, including 24 shared, 34 exclusive to 5ULTRA, and 14 to CADD. Removing 5ULTRA-annotated variants eliminated 15 of 38 CADD associations, indicating that a fraction of CADD’s performance depends on uORF and Kozak architecture. The 18 associations absent from a published UK Biobank 5′UTR study included 11 that were exclusive to 5ULTRA. Among these, *NELFCD*, a subunit of the RNA polymerase II pausing complex with no established role in megakaryopoiesis, reached −log₁₀*P* = 50 for platelet distribution width. Associations were most often driven by variants predicted to suppress translation (13 of 16 directionally resolved associations; *P* = 0.021). Gene-level effects of 5′UTR repressor variants correlated with those of coding protein-truncating variants (r = 0.68, *P* = 3.7 × 10⁻⁴), placing them on the same phenotypic scale. Associations tested in non-European participants showed 89% directional concordance (r = 0.84), supporting shared regulatory effects across ancestries. Mechanistic 5′UTR annotation therefore recovers a translational layer of phenotypic variation that generic deleteriousness scores based on evolutionary constraints miss.

## INTRODUCTION

The 5′ untranslated region (5′UTR) exerts control over protein output disproportionate to its length.^1^ Upstream open reading frames (uORFs) are short ORFs located 5′ to the primary coding sequence that can divert scanning ribosomes, induce stalling, or license downstream reinitiation, these effects together suppressing and regulating translation of the main ORF.^2^ More than half of human mRNAs carry at least one uORF,^3^ and massively parallel reporter assays have demonstrated that single-nucleotide variants disrupting uORF architecture can modify translational efficiency by one to two orders of magnitude.^4^ In addition to uORFs, the Kozak sequence context surrounding the main AUG determines ribosomal recognition efficiency; variants that strengthen or weaken this context can have phenotypic consequences.^5^ The 5′UTR is, thus, a densely encoded regulatory layer whose perturbation is increasingly linked to Mendelian disease, with pathogenic uORF-creating variants identified in various genes, including *GATA1*,^6^ *POLG*,^7^ and *BRCA2*^8^. The biological importance of the 5′UTR has been clearly established but this region has been little investigated in population-scale genetic studies.^8^ The recent completion of whole-genome sequencing for 490,640 participants in the UK Biobank project has transformed the study of 5′UTR variation. The dataset generated has revealed that 69.2% of 5′UTR variants are entirely missed by whole-exome sequencing,^9^ both highlighting the extent to which regulatory variation was previously inaccessible and providing a unique opportunity to study 5′UTR biology at population scale.

Despite their regulatory importance and the recent increase in their accessibility, there is a systematic underweighting of rare 5′UTR variants in population-scale association studies, for two different but compounding reasons. First, there is a problem with the score used. Predictors such as CADD^10^, a widely used pathogenicity predictor, integrates diverse genomic annotations but its assessment of noncoding variants is highly dependent on evolutionary constraint. It also lacks an explicit model of uORF-mediated translational regulation. As a result, 5′UTR variants that disrupt translation may be assigned moderate scores despite having substantial functional consequences. Some uORFs are lineage-specific^11^, arising and disappearing over short evolutionary timescales. A variant creating a strong new uORF in a human gene may, therefore, have a low score for evolutionary constraint simply due to variability of the corresponding position across vertebrates. Conversely, a position under strong constraint may harbor a variant that has no effect on uORF architecture. The second problem is one of directionality. Due to the mechanistic heterogeneity of 5′UTR variants, their aggregation into a single gene-level burden mask fundamentally conflates opposing biological effects. A uStart-gain variant creates a competing uORF that sequesters ribosomes and reduces the translation rate of the principal ORF, whereas a uStart-loss variant eliminates such an uORF and releases ribosomal flux towards the main ORF. A uStop-loss variant extending a non-overlapping uORF into the main ORF frame blocks reinitiation, whereas a uStop-gain variant truncating that same uORF restores reinitiation. The treatment of these mechanistically opposite variants as equivalent within the same burden test dilutes the signals from both directions simultaneously, resulting in a systematic loss of power that cannot be overcome by increasing sample size.

We have previously developed mechanism-specific genome-wide tools for other noncoding classes^12,13^ applying the same strategy to the 5′UTR, we developed 5ULTRA,^14^ a machine-learning classifier trained on six mechanistically different 5′UTR consequence classes (uStart-gain/loss, uStop-gain/loss, uKozak-change, and mKozak-change). 5ULTRA assigns functional scores to each variant by combining 17 uORF context features, including Kozak strength, uORF length, uORF-CDS overlap geometry, gene tolerance to variation, and uORF conservation across species.

The directional structure of these classes can be used to separate variants into two groups — those predicted to suppress CDS translation and those predicted to enhance it — making it possible to perform bidirectional burden tests that treat uORF gain-of-repression and loss-of-repression as distinctive biological signals rather than as noise, without each signal canceling the other out. In a previous GWAS of Mtb resistance, 5ULTRA contributed to the interpretation of a common-variant association signal and the prioritization of *YEATS4* as the candidate causal gene^15^. However, its potential for rare-variant gene-based association analyses, where functional annotation can be used to aggregate individually underpowered variants, has not yet been investigated. In this study, we applied 5ULTRA to data from 408,423 non-Finnish European participants from the UK Biobank WGS resource, to perform a genome-wide burden study of variants below 5% frequency, across 59 quantitative hematological and biochemical phenotypes. We addressed three questions: (1) Does mechanistic 5′UTR scoring increase gene discovery rates relative to CADD score in head-to-head benchmarking on the same cohort? (2) Do the opposing translational effects of UP and DN variant classes produce a detectable bidirectional translational dosage signal in population-scale association data? (3) Are 5′UTR loss-of-function burden effects quantitatively similar to exome protein-truncating variant effects for assessing clinical relevance? We obtained strong evidence for all three, showing that mechanistic 5′UTR annotation is a productive, synergistic complement to exome-centric rare-variant analysis.

## MATERIALS AND METHODS

### Study cohort

All analyses were performed under UK Biobank application 98772. We restricted the analysis to non-Finnish European (NFE) participants for whom whole-genome sequencing (WGS) data were available in the UKB 500K WGS release, accessed via the UK Biobank Research Analysis Platform (DNA nexus RAP; https://ukbiobank.dnanexus.com/landing). WGS data were extracted, by chromosome, from the ML-corrected DRAGEN population-level pVCF release, with bcftools view^16^ restricted to MANE Select 5′UTR intervals.^17^ The data were then merged across blocks, filtered to PASS variants with a genotype missingness < 10%, sorted, and converted to PLINK2 PGEN format with plink2.^18^ NFE ancestry was assigned according to the UK Biobank genetic ethnic grouping field (UKB field p22006), which identifies participants of white British and related European ancestry based on a genetic PCA performed by the UKB. After the exclusion of first-degree relatives (KING kinship coefficient > 0.0442)^19^ and individuals failing WGS quality control, 409,205 individuals were eligible for inclusion; complete phenotypic and covariate data were available for 408,423 of these individuals, who were included in the primary analysis.

### Phenotype selection and transformation

We used the 59 quantitative phenotypes from the UKB blood biochemistry (UKB field codes p30600–p30890, 30 traits) and complete blood count (p30000–p30300, 29 traits) panels, spanning hematological blood counts (platelet count, platelet crit, platelet distribution width, RBC count, MCH, MCV, MCHC, reticulocyte indices, lymphocyte and monocyte counts and percentages, eosinophil count), biochemical traits (HDL cholesterol, albumin, total protein, triglycerides, urate, alkaline phosphatase, ALT, oestradiol, HbA1c, cystatin C), and related determinations. Three considerations motivated this panel. First, both panels are assayed in every participant, so each trait is tested at full sample size. Second, rare 5′UTR burden signal concentrates in quantitative traits: in the UKB consortium’s own 5′UTR phenome-wide scan, 62 of 63 significant associations were quantitative and only one was a disease code.^9^ Third, published exome protein-truncating statistics exist for these same traits, allowing translational and coding effects to be compared on a common scale. Table S1 lists all 59 traits with their UK Biobank field IDs, the abbreviations used in the figures, and their full names. The hematology/biochemistry split follows the two UK Biobank assay panels (Blood count and Blood biochemistry); the finer organ-system labels within biochemistry (Figure 2) are descriptive only and do not correspond to UK Biobank categories. Before transformation, values more than five standard deviations from the cohort trait mean were set to missing. Phenotype values were then rank-based inverse-normal transformed (RINT) within REGENIE at association testing (--apply-rint).^20^ Covariates included age, age², sex, WGS sequencing batch, UK Biobank recruitment center, and the top 20 genomic principal components (25 covariates total).

### Variant annotation

Rare 5′UTR variants were annotated with the 5ULTRA pipeline^14^ (https://github.com/casanova-lab/5ULTRA). Starting from 5′UTR coordinates derived from MANE Select transcripts,^17^ we extracted all variants with a MAF < 5% and classified them into six consequence classes: uStart-gain, uStart-loss, uStop-gain (shorter non-overlapping; to non-overlapping), uStop-loss (longer non-overlapping; to overlapping), uKozak-change, and mKozak-change. Functional scores (range 0–1) were assigned to each variant by 5ULTRA. mKozak variants were assigned a fixed 5ULTRA score of 0.9999, reflecting their strong, context-independent effect on CDS Kozak context. Variants falling within annotated 5′UTR intervals but not annotated by 5ULTRA (those lying outside any uORF or Kozak regulatory context and not creating a new uORF) were assigned a default 5ULTRA score of 0.1 rather than 0, because a 5ULTRA score approaching 0 constitutes an active prediction of an absence of functional effect, whereas unannotated variants are of unknown significance and may influence translation, mRNA stability or transcription through unmodeled mechanisms. To derive burden weights, 5ULTRA scores were transformed to a PHRED-like scale (5ULTRA_PHRED = −10 × log_10_ (1 − 5ULTRA score)). This transformation increases the contribution of variants with the highest predicted functional impact while keeping low-scoring variants close to zero, thereby emphasizing variants most likely to affect translational regulation. CADD v1.7 scores^10^ were obtained from precomputed score databases. We further classified variants according to the direction of their effect on translation predicted by 5ULTRA: UP variants (uStart-loss; uStop-gain shorter non-overlapping; uStop-gain to non-overlapping) are predicted to increase CDS translation by reducing ribosome diversion; DN variants (uStart-gain; uStop-loss longer non-overlapping; uStop-loss to overlapping) are predicted to decrease CDS translation.

### Burden test design

Gene-level rare-variant burden tests were performed with REGENIE v4^20^ (--step 2, --qt, --build-mask sum, --apply-rint, --minMAC 0.5, --bsize 400, 16 threads). Step 1 whole-genome ridge regression predictions were calculated from the data of 408,423 NFE participants, with 727,178 LD-pruned common variants from the UKB imputed genotype array. Eight burden models were defined with two frequency thresholds (rare: MAF < 0.1%; af5: MAF < 5%): (1) **5ULTRA All**, including all 5′UTR variants with continuous 5ULTRA_PHRED weights; (2) **5ULTRA High-confidence,** restricted to variants with scores ≥ 0.5 retaining their continuous 5ULTRA _PHRED weights; (3) **5ULTRA UP or DN**, comprising separate UP and DN burden masks restricted to variants with an unambiguous directional consequence call, weighted by continuous 5ULTRA PHRED weights, with no score threshold; (4) **5ULTRA Joint (ACAT)**, combining the UP and DN burden test *p*-values with ACAT.^21^ (5) **CADD All**, including all variants of the MANE Select 5′UTRs weighted by continuous CADD scores; (6) **CADD High-confidence**, restricted to variants with CADD scores ≥ 5.0, weighted by continuous CADD scores (7) **CADD Attributable-Fraction High-confidence**, identical to the CADD High confidence mask except that all variants assigned a 5ULTRA consequence were excluded, and (8) **CADD Attributable-Fraction All,** identical to the CADD All mask after the exclusion of variants assigned a 5ULTRA consequence. The CADD Attributable-Fraction models quantify the fraction of the CADD association signal driven by variants falling within the scope of 5ULTRA annotation, as opposed to variants in 5′UTR regions not overlapping uORFs or Kozak regulatory elements. An additional, unweighted 5ULTRA DN model (af5, score ≥ 0.5, every qualifying variant given an equal weight of 1) was run for comparison with exome protein-truncating variant (PTV) burdens, which are themselves unweighted, to ensure that both were on the same scale. The 5ULTRA mechanistic suite comprises models 1-4; the CADD baseline suite comprises models 5 and 6.

### Burden quantification and statistical filtering

Burden magnitude was quantified as the aggregate weighted allele count (k) for each gene-phenotype-model combination. Each mask was built as a weighted sum of allele dosages (REGENIE --build-mask sum --weights-col 4), so k represents the sum of weighted allele contributions across all individuals rather than a raw carrier count. Gene-phenotype combinations for which k < 5 were excluded before association testing. All analyses were restricted to protein-coding genes with a MANE Select transcript. Across 59 phenotypes, eight 5ULTRA and four CADD model × spectrum combinations per gene, and after application of the k ≥ 5 filter, we performed 518,380 association tests in the 5ULTRA suite and 107,033 in the CADD suite. Genome-wide significance was therefore determined with Bonferroni correction for the exact number of tests performed in each suite: *P* < 0.05/518,380 = 9.65 × 10⁻⁸ (−log₁₀P > 7.016) for 5ULTRA and *P* < 0.05/107,033 = 4.67 × 10⁻⁷ (−log₁₀*P* > 6.331) for CADD. A nominal threshold of −log₁₀*P* ≥ 3.0 was applied for the 5ULTRA directionality analysis.

### 5ULTRA direction-of-effect analysis

For the 1,375 gene-phenotype pairs informative for directional analysis (k_UP ≥ 5 and k_DN ≥ 5, af5), the directional model gave separate burden estimates for variants predicted to decrease (DN, repressor) and increase (UP, enhancer) translation. Pairs were classified on the significance of each arm rather than on the sign of the coefficients: rheostat (opposite signs, both arms significant), DN-driven or UP-driven (only one arm significant, regardless of sign), genetic fragility (same sign, both arms significant), and class-nonspecific (UP and DN *P* > 0.05). Each arm was assessed at a nominal α risk of 0.05; this census is descriptive and uncorrected, and we report the number of pairs significant after Bonferroni correction separately. For the purposes of population-level description only, we recorded the proportion of pairs with *β*_DN × *β*_UP < 0. We compared this proportion with the 50% null hypothesis in an exact binomial test, and tested for a monotonic increase across significance bins with a Cochran-Armitage trend test. This descriptor has no significance gate and supports no per-pair claims of opposing effects. For the highlighted pairs, we refitted the repressor and enhancer burdens together in one model, in Python, according to RINT phenotype and with the covariates used in the REGENIE analysis (y ∼ covariates + burden_UP + burden_DN). Wald tests were then performed to determine whether the two effects were of the same size (β_UP = β_DN) and whether they were of the same size but opposite in sign (β_UP = −β_DN). This model does not build the burden in the same way as REGENIE: the burden is not capped, the phenotype is normalized before rather than after adjusting for covariates, and the whole-genome prediction from REGENIE Step 1 is not subtracted. Its coefficients are therefore on a scale different from REGENIE effect sizes, and we do not report them as effect sizes. The two tests are unaffected, because multiplying both coefficients by the same factor does not change whether they are equal. A rejected test means that the two effects differ in size, not that they act in opposite directions.

### Clinical equivalence comparison

Gene-level association statistics for coding protein-truncating variant (PTV) and missense variants were retrieved from the results published by the UK Biobank WGS consortium, the same source used for the 5′UTR study comparison above. For each gene-phenotype pair, the consortium reports a PTV burden effect and the single best-performing coding model. If the best model was one of their damaging-missense or nonsynonymous masks, its effect was used as a missense comparator, with the consortium’s own mask definitions and selection retained. The 5′UTR side used the unweighted 5ULTRA DN mask (af5, score ≥ 0.5; see Burden test design), matched with consortium statistics by gene and phenotype. Pairs were required to have a 5′UTR-DN carrier count k ≥ 15, with this floor value selected to ensure that the effect size of each pair was legible in Figure 4b. Correlations are shown for all carrier floors in Table S2. We performed Pearson correlation analyses of the relationship between 5′UTR and exome PTV Z-scores with the 5′UTR-UP (enhancer) mask, which does not model loss-of-function, as a negative control.

### Overlap with published 5′UTR associations

We intersected our significant genome-wide 5ULTRA results with associations reported in a previous UK Biobank consortium study of 5′UTR variants. The catalog comprised 42 gene-phenotype associations at −log_10_*P* ≥ 8.0. An association was considered consistent with the consortium report if the 5ULTRA burden effect was directionally concordant and significant under matching inclusion criteria (no minimum-carrier filter) and after Bonferroni correction (*P* < 0.05/42, −log_10_*P* > 2.92). A nominal significance threshold (*P* < 0.05, −log_10_*P* > 1.30) was also used as a sensitivity tier. For consistent associations, we compared the strength of the 5ULTRA association with the consortium result, classifying the signal as stronger (≥1.1×), similar (0.8– 1.1×), or weaker (<0.8×). We defined novel associations as significant genome-wide 5ULTRA gene-phenotype pairs that were absent from the consortium catalog, irrespective of the consortium significance level.

### Cross-ancestry generalization

The 58 significant genome-wide 5ULTRA associations detected in the NFE population (best model per gene-phenotype, −log₁₀*P* > 7.016, k ≥ 5) were evaluated for cross-ancestry generalizability in the UK Biobank non-European ancestry groups, with the same burden mask. The non-European cohorts comprised African (AFR, *n* = 8,048), South Asian (SAS, *n* = 9,871), East Asian (EAS, *n* = 1,569) and other/admixed (OTH, *n* = 73,242) participants. Associations were tested separately within each ancestry group and in a pooled non-European cohort (*n* = 92,730). In each ancestry group, only gene-phenotype pairs with values of k ≥ 5 were evaluated. Cross-ancestry generalization was assessed at three levels: (i) concordant effect direction; (ii) concordant effect direction and nominal *P* < 0.05; and (iii) concordant effect direction and Bonferroni-corrected *P* < 0.05/58. “Any ancestry” indicates that the association satisfied the corresponding criterion in at least one non-European ancestry group. No gene-phenotype pair reached k ≥ 5 in the South Asian or East Asian cohorts, so neither could be evaluated separately. This reflects the rarity or absence of the underlying variants in these populations.

### Data and code availability

5ULTRA software is available from https://github.com/casanova-lab/5ULTRA. The analysis code deposited at https://github.com/mchaldebas/5UTR-burden-UKB-release covers all steps downstream from REGENIE Step 2 and contains no individual-level data; all analyses of individual-level UK Biobank data must be performed on the UK Biobank Research Analysis Platform. UK Biobank data are available to approved researchers (www.ukbiobank.ac.uk). This research was performed with the UK Biobank Resource under application number 98772.

## RESULTS

### Evolutionary constraint on 5’UTR

We applied 5ULTRA to all 5′UTR variants in the UK Biobank WGS resource across six genetic ancestry groups (AFR, ASJ, EAS, NFE, SAS, OTH). This analysis yielded a mechanistically annotated catalog of 159,885 unique variants classified into six consequence classes (Figure 1a). The most abundant class was uStart-gain (62,559 variants), reflecting the frequency with which rare variants create *de novo* upstream AUGs in transcribed 5′UTR sequences; uStart-loss (36,891), uStop-loss (24,718), and uStop-gain (23,239) were the next most frequent classes, followed by uKozak-change (8,067) and mKozak-change (4,411). Across all classes, 5ULTRA functional scores were inversely associated with allele frequency in the NFE cohort: high-scoring variants (score ≥ 0.5, *n* = 42,521) had a mean allele frequency 5.1-fold lower than that of low-scoring variants (5.21 x 10^-4^ vs. 2.64 x 10^-3^; Mann-Whitney *P* = 3.7 x 10^-29^; Figure 1b). Variants in the highest score decile had a mean allele frequency 8.2-fold lower than those in the lowest decile, and a gradient was observed consistent with purifying selection acting preferentially on predicted functional variation. This inverse relationship between mechanistic impact score and population frequency provides orthogonal validation that 5ULTRA captures variation with biological consequences: the variants considered to have a high mechanistic impact are precisely those that evolution has been removing. Allele frequency distributions had consistent shapes across all six genetic ancestry groups for each consequence class, with the characteristic mass of ultra-rare singletons (AF ≈ 10^-6^) visible in all ancestries, confirming that the catalog is not dominated by any single population (Figure 1c).

**Figure 1.**
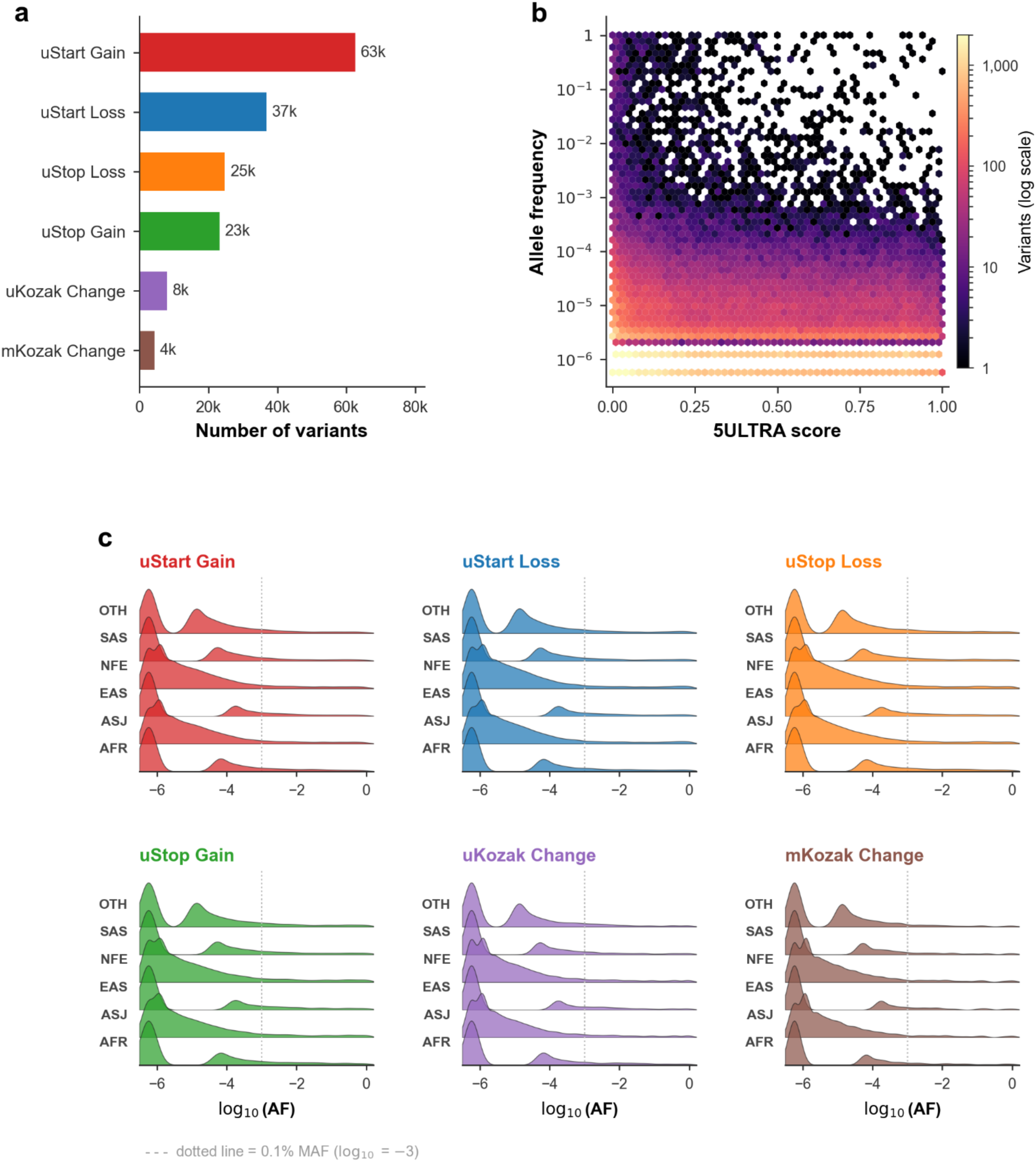
Global mechanistic landscape of 5′UTR variation in the UK Biobank WGS resource. (a) Horizontal bar chart of unique 5′UTR variant counts per consequence class across all six ancestral populations (*n* = 159,885 total). uStart-gain is the most abundant class (62,559 variants), reflecting the high frequency at which rare single-nucleotide variants create *de novo* upstream AUGs. Bar colors match the mechanism palette used throughout. (b) Hexbin density map of 5ULTRA score versus allele frequency for 114,414 NFE 5′UTR variants with AF > 0. The intensity of the color (log scale) indicates variant density per hexagonal bin. High-scoring variants (score ≥ 0.5) are concentrated at the lowest allele frequencies, with a mean AF 5.1-fold lower than that of low-scoring variants (Mann-Whitney *P* = 3.7 x 10^-29^), consistent with purifying selection acting on mechanistically impactful 5′UTR variation. (c) Ridge-line allele frequency distributions for each consequence class (columns) across six ancestral populations (rows). Square root-scaled kernel density estimates are plotted on the *y*-axis to enhance visibility; log_10_(AF) is plotted on the *x*-axis. The dotted vertical line corresponds to 0.1% MAF (log_10_(AF) = −3). The six consequence classes have near-identical shapes within each ancestry, with the bulk of variants at singleton frequencies (AF ≈ 10^-6^), confirming that mechanistic class membership does not introduce ancestry-specific allele frequency biases.

### 5ULTRA mechanistic scoring substantially expands 5’UTR rare-variant discovery

We tested the 59 blood-derived biomarkers (29 hematological indices and 30 serum biochemistry determinations) in 408,423 NFE participants, with rank-inverse-normal transformation of each of the values obtained (Table S1). The 5ULTRA models identified 58 significant genome-wide gene-phenotype associations (k ≥ 5, −log₁₀*P* > 7.0; Table S3), whereas the CADD models identified 38 such associations (k ≥ 5, −log₁₀*P* > 6.3; Table S4). We found that 24 of these associations were common to the two methods (9 genes, 19 traits), 34 were exclusive to 5ULTRA (16 genes, 24 traits), and 14 were exclusive to CADD (9 genes, 13 traits; Figure 2a). Hematological traits carried most associations (46 of 58 for 5ULTRA, 29 of 38 for CADD), but they were over-represented specifically among the 5ULTRA-exclusive discoveries. Hematological indices accounted for 29/59 traits tested (49%) but 26/34 5ULTRA-exclusive associations (77%; exact binomial P = 1.6 × 10⁻³). By contrast, the number of CADD-exclusive associations reflected the composition of the panel (9 of 14, 64%; P = 0.29). For 32 of the 34 associations that are exclusive to 5ULTRA, no CADD mask reached the k ≥ 5 testing threshold, so CADD returned no result rather than a null result. The remaining two associations were tested with CADD but were not significant (−log₁₀*P* 4.6 and 5.3). The clearest 5ULTRA-exclusive associations were those for *NELFCD* (platelet distribution width, −log₁₀*P* = 50), *CBFA2T3* (MCH, 31) and *APOA1* (apolipoprotein A, 29), for which the CADD masks contained essentially no qualifying variants (k ≤ 1; Figure 2b). The asymmetry ran both ways: 10/14 CADD-exclusive associations were not tested by 5ULTRA, so the two scores prioritize largely non-overlapping variant sets. Both scores were well calibrated on the high-confidence masks (AF < 5%): λ_GC_ = 1.01 for 5ULTRA (125,754 tests) and 1.05 for CADD (62,777 tests), with per-trait values of about one (Figure S1, Table S5). We then ablated all 5ULTRA-annotated variants from the CADD models: the number of significant genome-wide associations identified by CADD decreased from 38 to 30 (Figure 2c), with 15 associations lost and seven becoming significant only after ablation. All 15 of the lost associations were recovered by 5ULTRA, with 13 of genome-wide significance and 2 nominally significant, confirming that the ablated signal lies within the scope of 5ULTRA annotation. The associations that persisted reflect 5′UTR positions outside the uORF and Kozak context not annotated by 5ULTRA.

**Figure 2.**
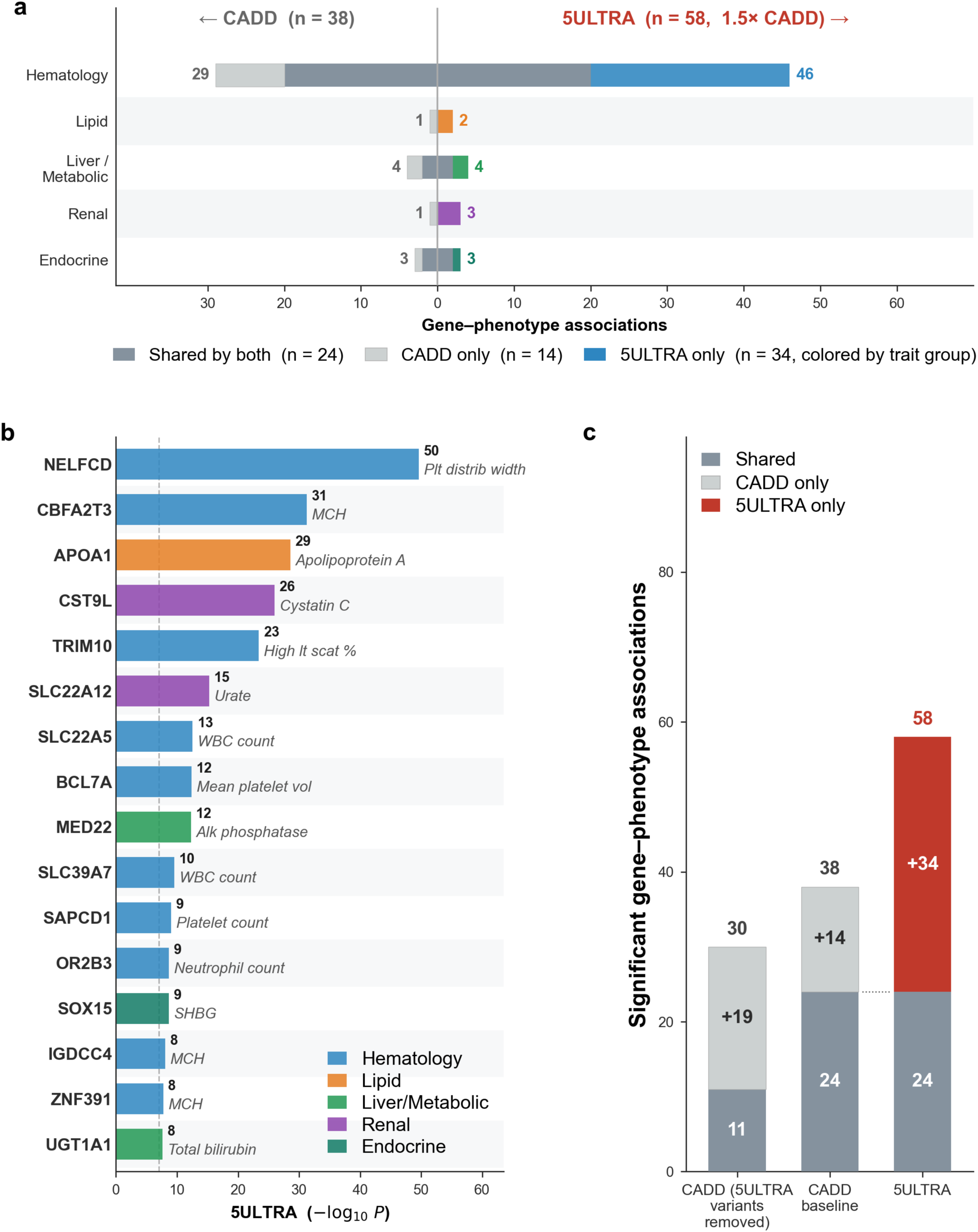
5ULTRA mechanistic scoring substantially expands rare-variant discovery across phenotype systems. (a) Butterfly chart of gene-phenotype associations at significance thresholds (5ULTRA: −log_10_*P* > 7.0; CADD: −log_10_*P* > 6.3; k ≥ 5), broken down into 5 phenotype groups. Thresholds are Bonferroni-corrected for the number of association tests performed in each suite (0.05/518,380 for 5ULTRA; 0.05/107,033 for CADD). Right side (colored bars): 5ULTRA associations; left side (gray bars): CADD associations. Shared associations (dark gray, from midline) represent gene-phenotype pairs detected by both tools; tool-exclusive associations (extending outward) show discoveries exclusive to the tool concerned. The numbers at the ends of the bars indicate the group total. Header labels show overall totals and the 1.5× fold-advantage of 5ULTRA over CADD. (b) Landscape of gene-phenotype associations exclusive to 5ULTRA (not detected by CADD at −log_10_*P* > 6.3), ordered by association strength. Each bar represents the top-scoring gene-phenotype pair for one 5ULTRA exclusive gene; bar color indicates phenotype group. Dotted vertical line marks the −log_10_*P* = 7.0 significance threshold. (c) CADD (5ULTRA variants removed): standard CADD burden models (High-confidence and All) with all 5ULTRA-annotated variants set to zero. CADD baseline: standard CADD burden models (High-confidence and All). 5ULTRA: mechanistic suite combining four models (All, High-confidence, UP/DN, Joint). Stacked bar segments: dark gray = common to both; red = exclusive to 5ULTRA; light gray = exclusive to CADD.

### 5ULTRA reproduces known associations and identifies new gene-phenotype pairs

We compared our results with the UK Biobank WGS Consortium Nature 2025 5′UTR PheWAS catalog (42 associations at −log₁₀P ≥ 8.0), applying the Consortium’s inclusion criteria (no minimum-carrier filter). Effect directions agreed for 30 of the 42 associations. Within these 30, the 5ULTRA burden reached nominal significance for 16 (P < 0.05; 38% of the catalog) and passed Bonferroni correction for 11 (0.05/42; Table S6). The 26 associations not reproduced therefore divide into 14 that were directionally concordant but fell short of significance and 12 whose effects pointed the other way. This is expected, as 5ULTRA is restricted to mechanistically annotated variants, so that its burden masks only partially overlap the Consortium’s all-5′UTR masks (Table S6). Consistent associations included those between *SLC4A1* and three erythrocyte indices (mean reticulocyte volume, −log_10_*P* = 15.9; mean sphered cell volume, 12.4; MCHC, 5.7) and *SOX15* (SHBG, −log_10_P = 10.9). 5ULTRA identified associations with 18 genes absent from the Consortium’s 5’UTR catalog (Table S7): *NELFCD* (platelet distribution width), *CBFA2T3* (MCH), *CST9L* (cystatin C), *TRIM10* (high light-scatter reticulocyte %), *MSTO1* (reticulocyte %), *MDC1* (RBC count), *BCL7A* (mean platelet volume), *MED22* (alkaline phosphatase), *SLC39A7* (WBC count), *BTNL2* (eosinophil count), *RPL7A* (alkaline phosphatase), *SAPCD1* (platelet count), *TRPM8* (total bilirubin), *OR2B3* (neutrophil count), *WDR46* (plateletcrit), *IGDCC4* (MCH), *ZNF391* (MCH), and *POM121L2* (MCH). 5ULTRA also yielded substantially higher power for five genes tested in the Consortium study but with results below its −log₁₀*P* ≥ 8.0 catalog threshold (Table S7): *SHBG* (SHBG), *GP1BA* (platelet distribution width), *APOA1* (apolipoprotein A), *SLC22A12* (urate), and *UGT1A1* (total bilirubin).

### 5′UTR associations usually involve repressor-class variants

The Directional model determines which functional class of variant is involved in an association. Across the 1,375 gene-phenotype pairs in which a gene has both repressor (DN) and enhancer (UP) variants (af5, k_UP ≥ 5 and k_DN ≥ 5), the two classes shifted the trait in opposite directions in 52.9% of cases (binomial *P* = 3.1 × 10⁻²; Figure 3a). This excess increased with signal strength, to 56% and 70% above −log₁₀*P* ≥ 2 and ≥ 3, although these subsets were small and not individually significant (Figure 3b). This approach considers direction without taking significance into account. We therefore also classified each pair according to the class that reached significance. At α = 0.05, 176 pairs (12.8%) displayed a directional signal, accounted for by a single class in almost all cases: 95 (6.9%) repressor-driven, 76 (5.5%) enhancer-driven, and 5 (0.4%) with both classes acting in the same direction. This analysis is restricted to genes carrying both classes of variant and excludes the strongest associations identified, in which enhancer variants were absent. After correcting for all 1,375 tests, one association remained, that of *BCL7A* with mean platelet volume. The discoveries displayed the same bias. Of the 58 significant genome-wide associations detected with 5ULTRA associations, 16 were best fitted by the Directional model and were therefore predominantly attributed to a single functional class: 13 were driven by translation-repressing variants and three by translation-enhancing variants (binomial P = 0.021; Table S3). Repressor variants carried the signal in eight of the 10 strongest pairs (Figure 3c), *TSEN2* across six traits, plus *OR2T6* and *SLC39A9*.

**Figure 3.**
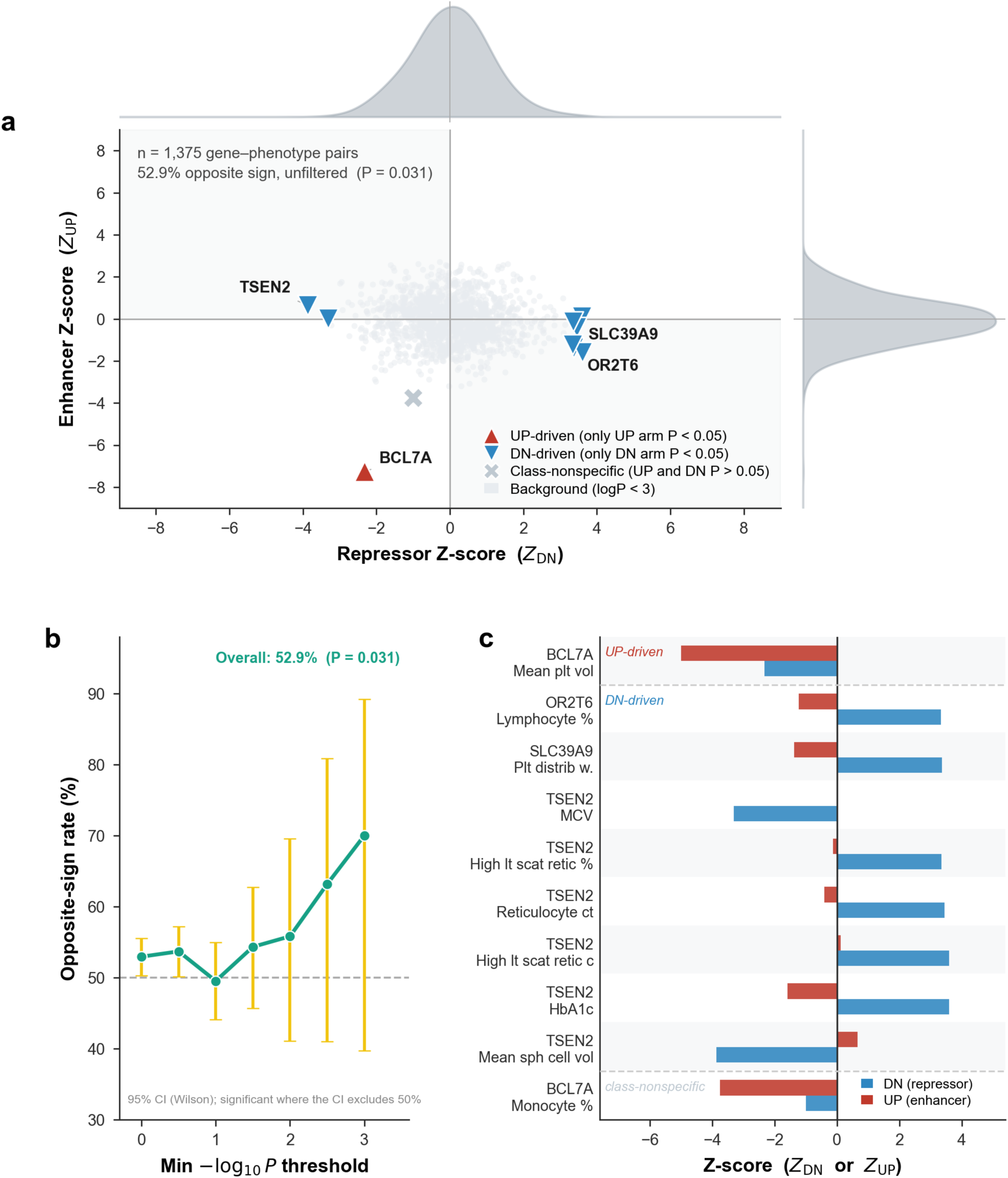
Strong 5′UTR burden associations involve predominantly repressor-class variants. (a) Repressor-class Z-scores (Z_DN, x-axis) versus enhancer-class Z-scores (Z_UP, y-axis) for all 1,375 gene-phenotype pairs with Directional-model results (af5, k_UP ≥ 5 and k_DN ≥ 5). Light gray points, background pairs (max −log₁₀*P* < 3.0). The ten pairs above this threshold are marked with the category assigned from the joint refit on the significance of each arm individually: blue downward triangles, DN-driven (only the repressor arm *P* < 0.05); red upward triangles, UP-driven (only the enhancer arm *P* < 0.05); gray crosses, class-nonspecific (UP and DN *P* > 0.05). Marginal kernel densities for all 1,375 pairs are shown above (Z_DN) and to the right (Z_UP). Across all pairs, with no significance filter, 52.9% of the pairs had opposite-signed coefficients (binomial *P* = 0.031 versus the 50% null hypothesis). (b) Proportion of pairs with opposite-signed coefficients above various −log₁₀*P* minima; dashed line, 50% null; gold bars, 95% Wilson confidence intervals for the proportion, with a threshold significant where its interval excludes 50%; only the two lowest thresholds (−log₁₀P ≥ 0 and ≥ 0.5) reach significance. (c) Repressor (DN, blue) and enhancer (UP, red) Z-scores for these ten pairs, grouped by category: UP-driven (BCL7A, mean platelet volume), DN-driven (OR2T6, SLC39A9 and TSEN2), and class-nonspecific (BCL7A, monocyte %).

### 5’UTR-DN burden effects are quantitatively similar to those of exome PTVs

Of the 359 gene-phenotype pairs reported with gene-level statistics for both coding protein-truncating variant (PTV) statistic and for which we had 5′UTR-DN variants, 23 passed the k ≥ 15 carrier floor. These pairs were not restricted to the 58 significant genome-wide associations. Across these 23 pairs, 5′UTR-DN burden Z-scores were significantly correlated with coding PTV Z-scores (Pearson r = 0.68, *P* = 3.7 × 10⁻⁴; Figure 4a), indicating that the phenotypic effects of 5′UTR repressor burden are comparable in magnitude to those of coding loss-of-function variants. The correlation did not depend on the carrier floor: it was significant at every threshold tested, from no k filter (*n* = 359, r = 0.34, *P* = 5.2 × 10⁻¹¹) to k ≥ 25 (*n* = 16, r = 0.70, *P* = 2.3 × 10⁻³; Table S2). This concordance was specific to the repressor direction: the enhancer (5′UTR-UP) burden was not correlated with exome PTV (r = 0.18, *P* = 0.62, n = 10; Figure 4a). Effect magnitudes, compared using the unweighted 5′UTR-DN mask for equivalence to the unweighted exome PTV burden, were directionally concordant for 19 of the 23 pairs (Figure 4b). At *GP1BA*, the 5′UTR-DN effect (+1.5 SD for platelet distribution width) exceeded the exome PTV effect (+0.4 SD), consistent with tight translational control of this platelet surface receptor. At *ALB*, the exome PTV effect (−2.5 SD) substantially exceeded the 5′UTR-DN effect (−0.1 SD), suggesting that albumin level is predominantly determined by coding sequence integrity rather than 5′UTR translational dosage.

**Figure 4.**
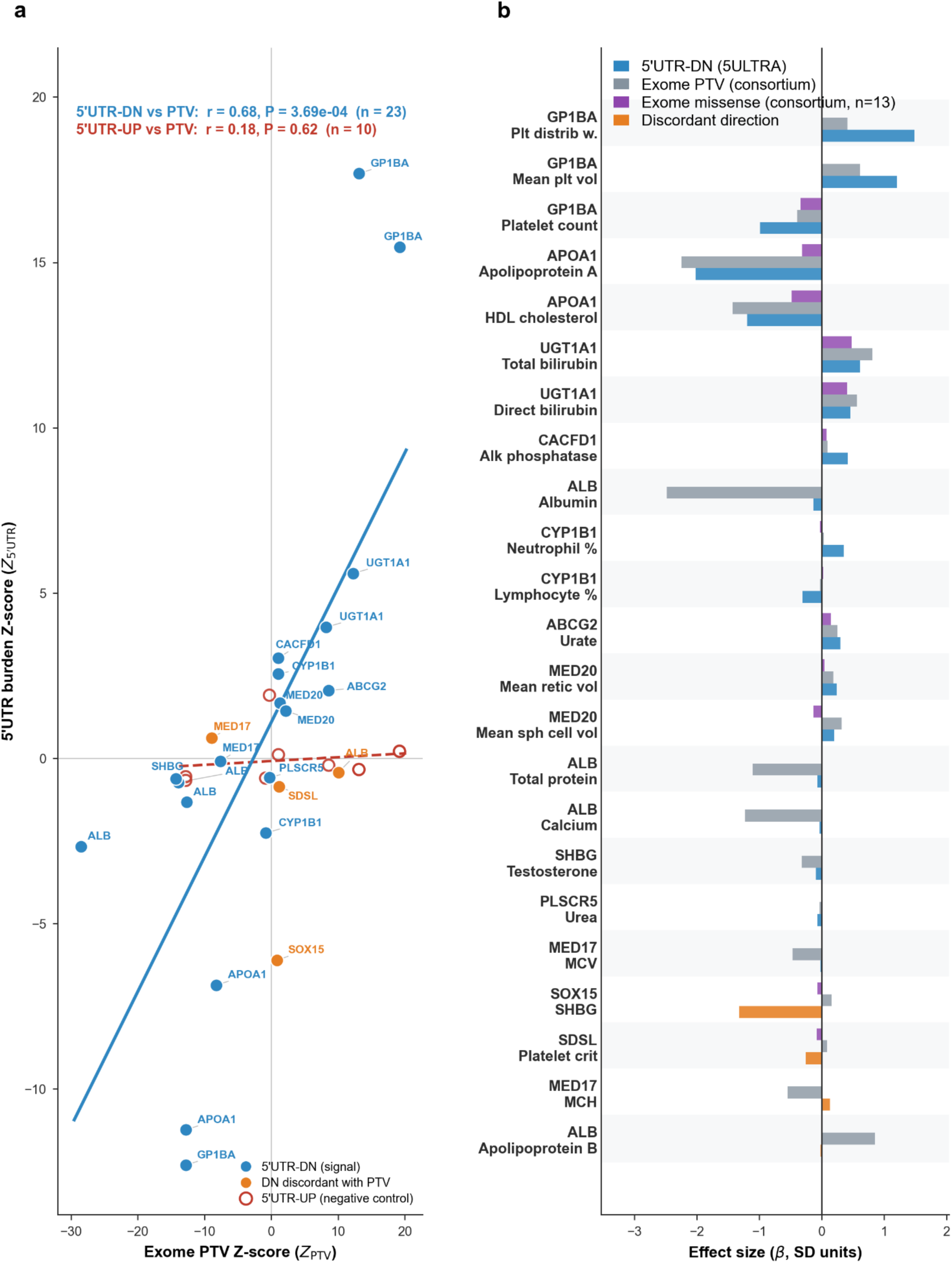
5′UTR-DN burden recapitulates exome PTV effects. (a) Exome PTV Z-score (*x*-axis) versus 5′UTR burden Z-score (*y*-axis) across gene-phenotype pairs (af5 spectrum, k ≥ 15). Closed circles, 5′UTR-DN (repressor) burden (unweighted 5ULTRA DN mask, score ≥ 0.5): blue = concordant with PTV direction, orange = discordant; solid blue line, DN regression. Open red circles, 5′UTR-UP (enhancer) burden (5ULTRA UP mask, af5) for the subset of pairs with UP data (*n* = 10), shown as a negative control; red dashed line, UP regression. (b) Per-pair effect sizes (β, SD units) for the 5′UTR-DN (blue/orange), exome PTV (gray), and best exome missense (purple; present for 13/23 pairs) masks, ordered first by concordance (pairs whose 5′UTR-DN and exome-PTV effects share the same sign, *β*DN × *β*PTV > 0, placed above discordant pairs), then by descending 5′UTR-DN burden −log₁₀*P*.

### Cross-ancestry directional concordance of associations

We investigated whether the associations discovered in the NFE cohort could be generalized beyond European ancestry, by retesting the 58 significant genome-wide 5ULTRA hits in 92,730 non-European UK Biobank participants, analyzed both within individual ancestry groups (African, n = 8,048; other/admixed, n = 73,242) and as a pooled non-European cohort (n = 92,730), using identical gene-phenotype burden masks. We found that 37 of the 58 significant genome-wide pairs were informative (k ≥ 5) in at least one non-European group (15 in AFR and 30 in OTH, 37 unique pairs); the others had too few carriers outside the NFE cohort, as expected for rare, often population-specific 5′UTR variants. When informative, effect directions were highly consistent: 33 of 37 (89%) were sign-concordant with the association discovered in the NFE population and non-European burden Z-scores were strongly correlated with NFE Z-scores in the pooled cohort (Pearson r = 0.84, *P* = 3.4×10⁻⁸, n = 27; Figure 5a). Formal validation was limited with the smaller non-European samples: 20 of 37 (54%) reached nominal significance (*P* < 0.05) and 9 of 37 (24%) survived Bonferroni correction across the 58 tests (Figure 5b). The African cohort illustrates this power limitation: 11 of 15 informative pairs were directionally concordant (73%) but none reached formal significance at this sample size. These results demonstrate substantial cross-ancestry concordance of 5ULTRA associations, supporting shared regulatory effects across ancestry groups while highlighting the limited power of the current non-European sample sizes for rare-variant association testing.

**Figure 5.**
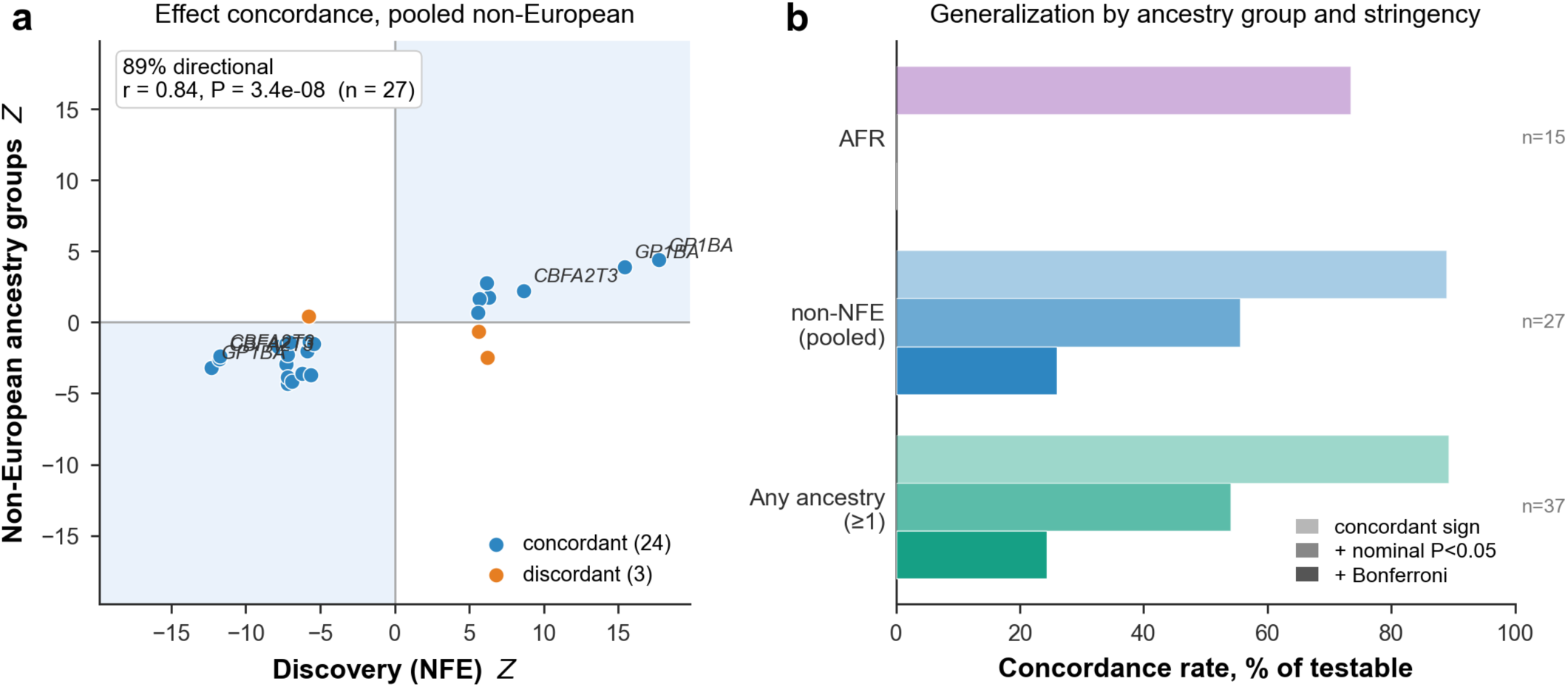
Cross-ancestry generalization of 5ULTRA associations. (a) Discovery (NFE) burden Z-score versus non-European burden Z-score for the significant genome-wide 5ULTRA hits testable in the pooled non-European cohort (*n* = 27). Blue, direction-concordant; orange, discordant; shaded quadrants indicate concordance; 24 of 27 pairs (89%) are directionally concordant (Pearson r = 0.84, *P* = 3.4×10⁻⁸). (b) Concordance rate (% of testable hits) in the African (AFR) ancestry group, the pooled non-European ancestry group, and across any non-European ancestry group (concordant in ≥ 1 non-European ancestry group), at three stringencies: concordant sign; concordant + nominal significance (*P* < 0.05); concordant + Bonferroni-corrected significance (*P* < 0.05/58). *n* = hits testable (k ≥ 5) in each group; no AFR hit reached nominal or Bonferroni significance.

## DISCUSSION

Most deleteriousness scores for non-coding variants are based principally on evolutionary constraint: they consider whether a position has been conserved rather than the molecular event caused by a variant. By reading the translational grammar of the 5′UTR directly (uORFs, Kozak context, and the initiation events they govern) 5ULTRA provides a mechanistic information absent from these other scores. Across 408,423 UK Biobank genomes and the 59 quantitative blood traits measured in every participant and therefore tested at full sample size, 5ULTRA burden models identified 58 significant genome-wide gene-phenotype associations, including 34 not recovered by CADD-based burden models. This substantial gain suggests that mechanistic annotation captures biologically relevant information beyond generic estimates of variant deleteriousness, thereby improving the sensitivity of rare variant association analyses while maintaining specificity. The complementary, rather than redundant, nature of 5ULTRA and CADD was illustrated by the ablation analysis. Removing 5ULTRA-annotated variants from the CADD burden models eliminated 15 of the 38 associations, all of which were recovered by 5ULTRA, indicating that a substantial proportion of the power of CADD to discover 5′UTR associations is based on variants affecting uORF and Kozak features. The residual CADD-only signal corresponds to layers of regulation — structured RNA, non-AUG initiation and transcriptional control — beyond the current six-class ontology, defining the next frontier. When tested against the concurrent UK Biobank Consortium catalog, 5ULTRA recovered 16 of the 42 reported associations when matched criteria were applied (11 after Bonferroni correction) and, more tellingly, identified 18 genes absent from the Consortium catalog. Mechanism, not conservation, is what these variants have in common. Association discovery was performed in the NFE cohort, but the associations were directionally consistent in non-European cohorts, indicating that this signal is not specific to individuals of European ancestry; larger non-European samples are now required for formal generalization.

The most consequential finding is not a longer list of genes but a principle: the 5′UTR acts as a directional, predominantly repressor-driven translational dosage layer. Variants that build or strengthen inhibitory uORFs (DN) decrease translation of the main ORF translation and behave like loss-of-function variants. Those that dismantle uORFs (UP) release ribosomal flux and behave like gain-of-function variants. If the trait considered is the amount of protein produced, then repressor and enhancer variants would be expected to push phenotypes in opposite directions. Our data are consistent with that expectation but do not formally demonstrate it: 52.9% of 1,375 informative pairs had opposite-signed coefficients, but significance in both arms was not achieved for any of the pairs, so no individual locus demonstrated opposition. What the discoveries do show is that where a directional signal is resolved, it is usually the repressor arm that carries the signal: 13 of the 16 significant genome-wide directional-model associations acted through the DN mask (binomial *P* = 0.021). *NELFCD*, the single most significant discovery (−log_10_*P* = 50), links translational control of a transcription-pausing factor to megakaryopoiesis, a connection that has never been reported. *TSEN2* had the clearest repressor-carried signal; its DN burden was associated with six traits whereas the enhancer arm was not independently significant. *BCL7A* is the informative exception: its association with mean platelet volume is carried by the enhancer arm.

Translationally repressive 5’UTR variation can produce phenotypic effects comparable in magnitude to coding loss-of-function. Across 23 matched genes, 5′UTR-DN burden tracked coding PTV burden with r = 0.68 (*P* = 3.7 × 10⁻⁴), placing the two in the same phenotypic units. However, the agreement is most informative when it breaks down. At *GP1BA*, the 5′UTR-DN effect on platelet distribution width dwarfs the heterozygous PTV effect (+1.5 versus +0.4 SD), potentially reflecting phenotypic sensitivity to gradual translation reduction that differs from the consequences of coding loss-of-function. At *ALB,* the pattern was inverted: the PTV effect substantially exceeded the 5′UTR-DN effect (−2.5 versus -0.1 SD), consistent with a larger phenotypic impact of coding loss-of-function than of 5′UTR-mediated translation reduction. Population-scale 5′UTR burden testing therefore does something that a generic deleteriousness score cannot: it provides an empirical comparison of the relative phenotypic consequences of translationally disruptive 5′UTR variation and coding loss-of-function at individual loci. These observations may help identify genes for which non-coding 5′UTR variation deserve particular attention, in rare-disease sequencing, in both research and clinical settings.

There are boundaries to these conclusions. The six-class ontology omits structured RNA^22^, internal ribosome entry sites^23^, m6A modification^24^, and non-AUG initiation^25^. We also restricted testing to variants below 5% frequency. Common variants in the UK Biobank have already been surveyed exhaustively, so 5ULTRA is unlikely to yield new associations there. Its contribution at that end of the spectrum is interpretive rather than exploratory, resolving which variant under a GWAS peak is causal. Bidirectionality is a genome-wide trend with limited power at any single locus, and the PTV comparison borrows from published statistics, the heterogeneity of which can only dilute, never inflate, the correlation reported. However, none of this undermines the central claim. Three independent lines of evidence — a 1.5-fold expansion of discovery, a directional architecture in which repressor-class variants carry the most clearly resolved signals, and a quantitative equivalence to coding sequence loss-of-function — converge on one conclusion: mechanistic annotation of the 5′UTR recovers a stratum of human phenotypic variation that generic deleteriousness scores do not take into account. A component of phenotypic variation currently attributed broadly to noncoding variation may therefore reflect altered translation at dosage-sensitive loci. Mechanistic scores predict not only the variants that really matter but also the direction in which they act, thereby convert population genetics into testable physiology. The 5′UTR provides the proof of principle that it is by focusing on mechanism, in addition to conservation, that rare-variant genetics will be able to find what it has been missing.

## Data Availability

https://github.com/mchaldebas/5UTR-burden-UKB-release

https://www.ukbiobank.ac.uk

## ACKNOWLEDGMENTS

The Laboratory of Human Genetics of Infectious Diseases was supported by the Howard Hughes Medical Institute, the University of Texas Southwestern Medical Center Governor’s University Research Initiative (GURI) grant program (04-2026), the French Agence Nationale de la Recherche (ANR) under the France 2030 program (ANR-10-IAHU-01), the Integrative Biology of Emerging Infectious Diseases Laboratory of Excellence (ANR-10-LABX-62-IBEID), the French Foundation for Medical Research (FRM) (EQU202503020018), the Square Foundation, Grandir - Fonds de solidarité pour l’enfance, the Fondation du Souffle, the SCOR Corporate Foundation for Science, the Battersea & Bowery Advisory Group, University of Texas Southwestern Medical Center, Institut National de la Santé et de la Recherche Médicale (INSERM), Paris Cité Université, and the Imagine Institute. H.M. was supported by the Eunice Kennedy Shriver National Institute of Child Health & Human Development of the National Institutes of Health under Award Number F30HD116571, and a NIGMS/NIH Medical Scientist Training Program grant T32GM152349 to the Weill Cornell/Rockefeller/Sloan Kettering Tri-Institutional MD-PhD Program. The content is solely the responsibility of the authors and does not necessarily represent the official views of the National Institutes of Health.

## WEB RESOURCES

5ULTRA, https://github.com/casanova-lab/5ULTRA

Analysis Code, https://github.com/mchaldebas/5UTR-burden-UKB-release

UK Biobank, https://www.ukbiobank.ac.uk

