## Supplemental Figure 1 for "Mechanistic 5’UTR Variant Scoring Expands Rare Variant Discovery in the UK Biobank"

### Supplemental Figures

**Figure S1: Genome-wide burden association landscape for 5ULTRA and CADD.**

(a) Miami plot of $-\log_{10}P$ values for all gene-phenotype burden tests (carrier count $k \geq 5$) across 59 quantitative traits in 408,423 NFE participants. Upper panel: 5ULTRA suite (best of 4 models × 2 frequency spectra per gene-phenotype pair); lower panel (inverted): CADD suite (best of 2 models × 2 spectra). Dashed red lines indicate genome-wide significance thresholds ($-\log_{10}P > 7.0$ for 5ULTRA; $-\log_{10}P > 6.3$ for CADD). Top associated genes are labelled with their peak $-\log_{10}P$. (b, c) Quantile-quantile plots of observed versus expected $-\log_{10}P$ for the 5ULTRA High-confidence af5 model (b) and CADD High-confidence af5 (c); points above the genome-wide significance threshold are highlighted. Genomic inflation factors $\lambda_{GC} = 1.011$ (5ULTRA) and $\lambda_{GC} = 1.045$ (CADD).

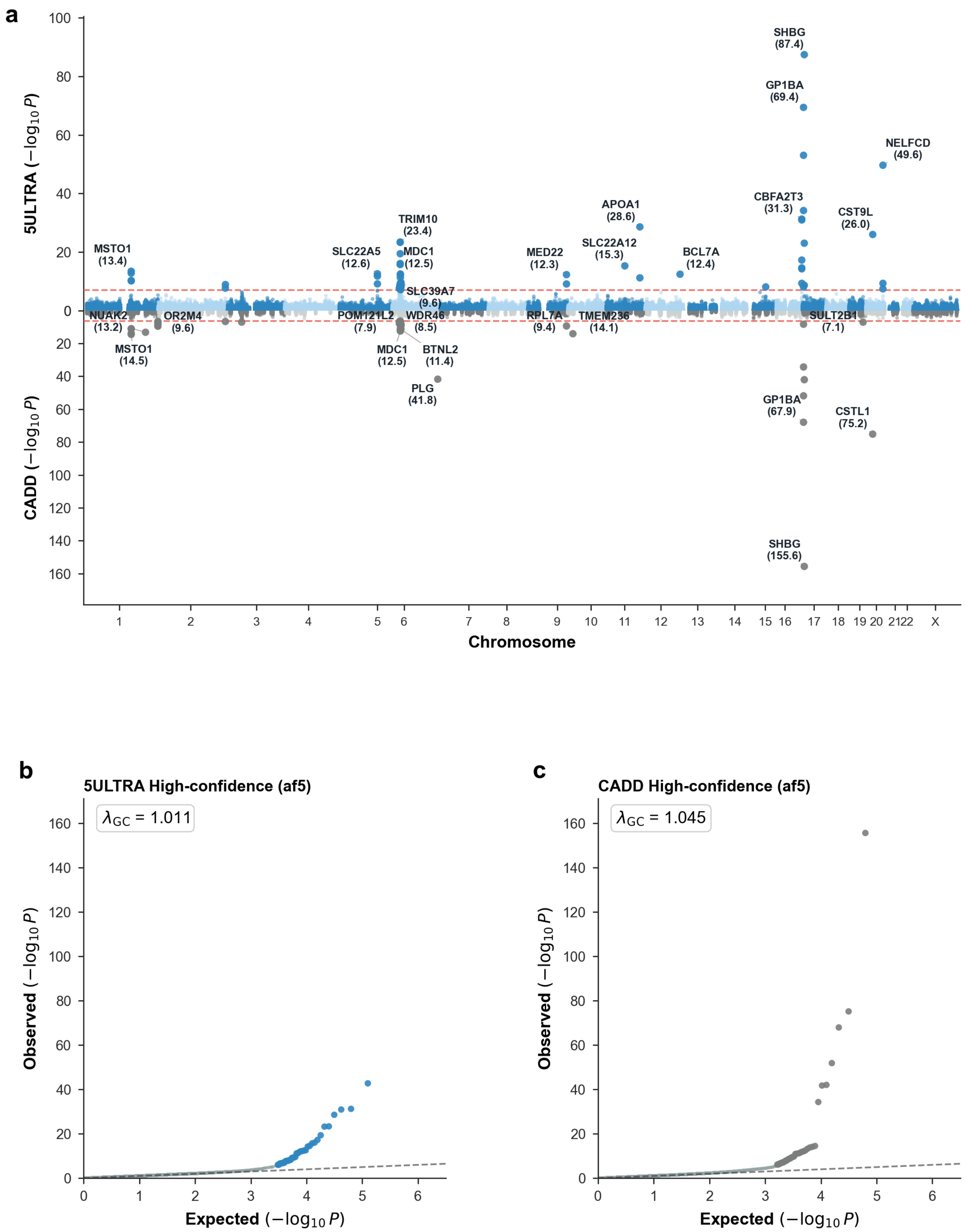
a
5ULTRA (−log10 P)
CADD (−log10 P)
Chromosome
SHBG (87.4)
GP1BA (69.4)
NELFCD (49.6)
CBFA2T3 (31.3)
CST9L (26.0)
APOA1 (28.6)
SLC22A12 (15.3)
BCL7A (12.4)
MED22 (12.3)
TRIM10 (23.4)
MDC1 (12.5)
SLC22A5 (12.6)
SLC39A7 (9.6)
MSTO1 (13.4)
NUAK2 (13.2)
MSTO1 (14.5)
OR2M4 (9.6)
POM121L2 (7.9)
WDR46 (8.5)
MDC1 (12.5)
BTNL2 (11.4)
PLG (41.8)
RPL7A (9.4)
TMEM236 (14.1)
SULT2B1 (7.1)
GP1BA (67.9)
CSTL1 (75.2)
SHBG (155.6)
b
5ULTRA High-confidence (af5)
λGC = 1.011
Observed (−log10 P)
Expected (−log10 P)
c
CADD High-confidence (af5)
λGC = 1.045
Observed (−log10 P)
Expected (−log10 P)
